# Well-being focused interventions for caregivers of children with developmental disabilities-a scoping review protocol

**DOI:** 10.1101/2022.06.29.22277042

**Authors:** Nokuthula Nkosi Mafutha, Doreen Asantewa Abeasi, Joseph Suglo

## Abstract

**Introduction:** Children with developmental disabilities (DD) have complex health needs which imply that they will need assistance in many areas of their lives, a role usually assumed by family members. Family caregivers need to constantly balance the demands of providing care with their own needs, this may be overwhelming and may lead to elevated levels of stress, subsequently affecting their well-being. For optimum functioning of the child with DD, the caregiver would have to be in a state of good health, making it necessary to have interventions that will support caregivers’ well-being. This scoping review aims to identify and map the range of interventions available for caregivers of children with DD.

**Methods and analysis:** This scoping review looks at well-being interventions of caregivers of children with DD. Two reviewers who are independent and blinded will search of articles from five databases, namely; CINAHL, Psych INFO, PubMed, ERIC and COCHRANE Library. Grey literature will be looked for from google scholar. Predetermined inclusion criteria will be used in the literature search. The findings will be summarized in a tabular form, followed by narrative text.

**Ethics and dissemination:** As a scoping review of published literature, ethics approval is not required. Results will be disseminated through publication in a peer-reviewed journal.

**Discussion:** The proposed scoping review will give an extensive review of interventions aimed at improving the well-being of caregivers of children with DD. We hope that this scoping review will provide recommendations on well-being improving interventions for caregivers of children with DD. Additionally, the review will guide future work on intervention development and primary research in this field.

**Registration:** This scoping review protocol has been registered with the Open Science Framework (https://osf.io/tkbrh)

**Strengths and limitations of this study:**

- This scoping review when completed will be the first study to identify and review systematically well-being focused interventions for caregivers of children with developmental disabilities.
- The scoping review will employ standard guidelines of Preferred Reporting Items for Systematic Reviews and Meta-analysis extension for scoping review (PRSMA-ScR).
- The search will cover a broad scope by using five databases as well as grey literature sources.
- Inter-rater reliability will be increased by at least two raters/reviewers doing abstract screening, full text screening and data extraction. A third reviewer will be included to resolve the disagreements which will arise from the other two reviewers.
- Quality assessment of studies included in the scoping review will be done.
- The scoping review will be limited to articles published in English and those from 2010 to 2021.

## Introduction

Globally, about a 100 million children under the age of 15 years are reported to have developmental disability (DD) ^1^. The estimated number of children aged 0-5 years with DD had risen from 598.5 million in 1990 to 632.0 million in 2016. Low- and middle-income countries (LMIC) accounted for the majority (50.2 million) with the remaining representing high-income countries (HIC) ^2^. Boyles et al. conducted a study on the trends in the prevalence of developmental disabilities in children from 3 to 17 years in the United States between 1997 and 2008 using a parent report diagnosis ^3^. The conclusions were that over the past 12 years there was an increase in DD from 12.84% to 15.4%. They concluded that DD were quite common in the US as approximately 1 in 6 had DD between 2006-2008. In a more recent study by Zablotsky et al. conducted between 2014 and 2016, the prevalence of DD of children between 3 to 17 had increased from 5.76% to 6.99% ^4^. Their study was not based on patient report as in Boyles et al’s study but rather on clinical diagnosis. Comparing the findings of Boyle et al’s to Zablotsky et al, the reported prevalence was lower in the latter study which the authors attributed to a more restricted definition of the concept of DD which they utilized ^3,4^.

Children with DD have long term multiple impairments and activity limitation as well as complex health care needs. They have problems with feeding, sleep and challenging behaviours. Difficulty in feeding which happens to be serious and lasting is associated with children with DD ^5^ than typically developing children. It is estimated that about 35% of children with some form of neurodevelopmental disabilities have feeding problems compared to only 25% in typically developing children ^6^. Children with DD suffer from various forms of sleep problems which is usually severe and enduring ^7^. The estimates vary across studies but there is a general consensus that the prevalence of sleep problems is higher in children with DD than typically developing children and the general population ^8^. Most common challenging behaviours include; aggression, hyperactivity, impulsivity, self-injurious behaviours and destructive behaviours ^9^.

The management of children with DD therefore requires ongoing care and management, and resources. Family members usually take up the responsibility of caring for a sick person, be it acute or chronic ^10^. Dealing with the needs of children with DD on long term basis are usually stressful as several studies have reported that caregivers of such children experience higher levels of stress ^11,12^ and increased burden of care ^13,14^. Subsequently, high stress levels negatively affect caregivers physical ^15^, psychological and social wellbeing ^16,17^. For example, chronic stress has been documented to negatively affect caregivers cardiovascular, immune and gastrointestinal systems ^18,19^ and this is linked to disturbances in the cortisol patterns ^20^. Somatic symptoms like headaches, backaches, gastrointestinal problems, respiratory infection have been reported ^21,22^ in caregivers of children with disabilities.

The challenge is that poor health status of the caregiver is linked to frequent hospitalizations of the child with DD ^23^, emotional and behavioural problems ^24^, lower health related quality of life of the child ^25^, negative parenting behaviour in which caregivers are irritable and hostile towards the child ^26^ accompanied by lower quality of supervision ^27^. This literature highlights the importance of the provision of effective support for caregivers for their own benefit and for the benefit of the children with DD. Scoping review done in this area have been limited to interventions for children with disabilities ^28,29^. Those scoping review that have looked at caregiver-related issues did not specifically consider interventions to improve their well-being but rather considered the burden of care, impact and needs of caregivers ^30^. Though, one study examined interventions for primary caregivers, the focus was on only one aspect of well-being and it was limited to caregivers of children with Autism Spectrum Disorders ^31^. Thus, no scoping review exists on the interventions done for caregivers of children with DD. In this scoping review, we aim to identify and evaluate existing interventions for improving caregiver well-being. We also aimed to identify gaps in the literature through evidence mapping.

## Review aims

The aim of this scoping review is to describe type, content, delivery and outcomes of interventions which focused on improving the well-being of caregivers of children with DD and identifying gaps in knowledge. The findings from the proposed scoping review could potentially be valuable to inform policy makers, clinicians and researchers on what needs to be done to improve the overall well-being of caregivers of children with developmental disabilities.

## Methods and analysis

This scoping review process will be guided by the methodological framework of Arksey and O’Malley as well as the Joanna Briggs Institutes (JBI) guidelines. The scoping review process will comprise of six stages; 1) identifying the research question; 2) identifying relevant study; 3) study selection; 4) charting the data; 5) collecting, summarizing and reporting the results and 6) a consultation exercise ^32–34^

### Stage 1: Identifying the research question

i. What types of interventions have been implemented and evaluated for caregivers of children with developmental disabilities?
ii. What are the characteristics of interventions for caregivers of children with developmental disabilities?

### Stage 2: Identifying relevant studies

The search is usually broad as it is not limited only to data from specific forms of evidence, thus, regardless of the research approach, be it qualitative or quantitative ^34^. The search is supposed to be comprehensive enough to identify other sources of primary studies (grey literature). JBI provide a three-step strategy and this will be utilized in the scoping review^34^. The first step will begin with limited search of at least two databases which are relevant to the study. The aim of this section will be to analyze text words that are found in the retrieved papers with the focus being on the title and the abstract. The key words or index terms will also be analyzed. The second step will involve conducting another search using all the index terms that will be identified in step one. In this case, the search will not be limited to only two databases but rather it will be done across all databases that is to be used in the scoping review. The final step will involve searching the reference list of all identified studies for additional studies. The key words and corresponding alternative words are shown in table 1.

**Table 1.** Search strategy.

| Key word | Alternative words |
| --- | --- |
| Caregiver | father, mother, parent, family, carer |
| Children | juvenile, young adult, teenager, adolescent |
| Developmental disability | developmental delay, cognitive disabilities, developmental disorders, intellectual disability, learning disabilities, neurodevelopmental disabilities, intellectual and developmental disabilities, cognitive impairment, mental retardation |
| Intervention | education, training, programme, therapy |
| Well-being | wellness, health |

#### Initial Search

Two databases (CINAHL Complete and Psych Info) will be used for the initial search. The search terms will be caregiver, carer, child, children, developmental disability, intellectual disability, developmental delay, intervention, programme, training, well-being, wellbeing, well being. Boolean operators like ‘OR’, ‘AND’ will be used to link the words for the search. The search will be limited to January 2010 to September 2021 and journal articles in English language.

#### Search across five databases

Five databases will be searched namely; PubMed, PSYCHINFO, Cochrane Library, ERIC and CINAHL complete. The following key words will be used; caregiver, well-being, interventions, child, and developmental disabilities. In a scoping review, it is always important to construct a comprehensive search with the aim of not missing out on any study or article. For the databases search, the indexing terms or medical subject headings (MeSH) specific to the particular database will be used. Apart from the MeSH or indexing term for a particular keyword, alternative keywords will be used. For example, a phrase like developmental disabilities, alternative key words such as developmental delay, cognitive disability or neurodevelopmental disability or mental retardation will be used. Additionally, to broaden the search, the root word will be used and truncation symbol will be put at the end. For example, famil* will yield results of family and families. Quotation marks will be used for phrase searching to ensure that the results retrieved from the search engine are accurate. A phrase like developmental disabilities will bring back results that have those words in them. For example, in PubMed a search returned 32, 208 results whilst “developmental disabilities” returned 28, 735.

The search in the databases will be done database specific. Boolean operators like ‘OR’, ‘AND’ will be used to link the words for the search. The final search strategy will be developed by the NN and a librarian at the University of Witwatersrand, Faculty of Health and Medical Sciences library.

As an illustration, we will input the following key words to search articles in the CINAHL database: (MH “Caregivers” OR father* OR mother* OR parent* OR famil* OR care-giver* OR caregiver* OR carer*) AND (MH “Wellness” OR “well-being” OR health) AND (MH “Psychosocial Intervention” OR educat* OR train* OR program* OR therap* OR intervention* OR treatment* OR skill*) AND (MH “Child” OR child* OR juvenile OR “young adult*” OR teenage* OR adolescen*) AND (MH “Developmental Disabilities” OR “developmental delay*” OR “cognitive disabilit*” OR “developmental disorder*” OR “intellectual disabilit*” OR “learning disabilit*” OR “neurodevelopmental disabilit*” OR “intellectual and developmental disabilit*” OR “cognitive impairment*” OR “mental* retard*”)

#### Searching the reference of identified studies/other sources

The reference list for identified studies will be searched to identify additional studies. This is important because the researchers will like to exhaust all possible means and not stand the chance of excluding some studies.

### Stage 3: Study screening and selection

Screening and selection of the studies will be guided by the predetermined inclusion and exclusion criteria. The results from each of the database will be imported to a reference manager, Mendeley to remove duplicates. After removal of the duplicates, the articles will be exported to Covidence, a review management system for screening of the articles. The screening of the titles and the abstract will be done independently by two reviewers (DAA and JNS) for eligibility. Differences which will arise among the reviewers will be resolved by NN. The screening of titles and abstracts will be followed by the full text review done independently by two reviewers (DAA and JNS). In the event of a disagreement, a third reviewer (NN) will make the final decision.

#### Review inclusion criteria/Eligibility criteria

The following criterion, will be applied in selecting the studies:

#### Population

Primary caregivers of children with DD who are 18 years and above and children with DD between the ages of 5 years and 14 years will be considered. Caregivers of children with DD who are less than 18 years, caregivers who are not the primary caregivers and adults with DD will not be included. Caregivers who may not be intensely involved in the care of the child with DD and be only providing a supporting role may have different health needs and experiences as compared to the primary caregiver. Adults with DD may have different needs as compared with children, as such, they will also be excluded.

#### Concept

We will include interventions which aim to improve the well-being of caregivers of children with developmental disabilities. Interventions not focused on the health of the caregiver and those on the child with DD will not be included. Interventions focused on the child with DD may not necessarily have an impact on the primary caregiver. Again, not all interventions are tailored towards the well-being of the caregiver.

#### Context

The context of the current review will not be restricted to a particular geographical location. Though the current study is being conducted at low resource setting and it would have been appropriate to consider studies form those settings, only a handful of studies have been conducted in these setting, giving the need to consider studies from all geographical areas.

#### Types of studies

Studies to be included in the scoping review are; Randomized Control Trial, Quasi-experimental (Cohort) Trial, Single Group Trial, Single Subject Experimental Design. Studies such as opinions. Conference presentations, dissertations, discussion papers, case study, book, editorials or commentaries and systematic review will be excluded. Interventions are aimed at assessing its impact on certain outcome variables. It will be impossible to achieve this with opinion papers and others. Though SR and MA are evidence-based, they are also pool of interventions put together and such evidence may not be relevant for the scoping review.

Languages other than English will be excluded as the researchers may need to translate those studies before they could be used. A few studies were conducted prior to 2010. For example, a search in PubMed without any restriction on the date of publication retrieved 778 results whilst a restriction yielded 482 results. Thus, studies published from January 2010 will be included.

#### Types of outcome measures

All types of interventions will be considered, including those that will focus on one particular outcome or multiple outcomes. Thus, interventions could address any or all of the following areas: stress, burden, social support, physical health.

### Stage 4: Data extraction

Charting the results is also referred to as extraction of results (JBI, 2015). Charting results provides a descriptive and logical summary of the results. It is an iterative process as the charting table is updated continually. The information that will be charted include; author(s), year of publication, origin/country of origin, purpose of study, study population and sample size, methodology, intervention type, and key findings and shown in table 2.

**Table 2:**
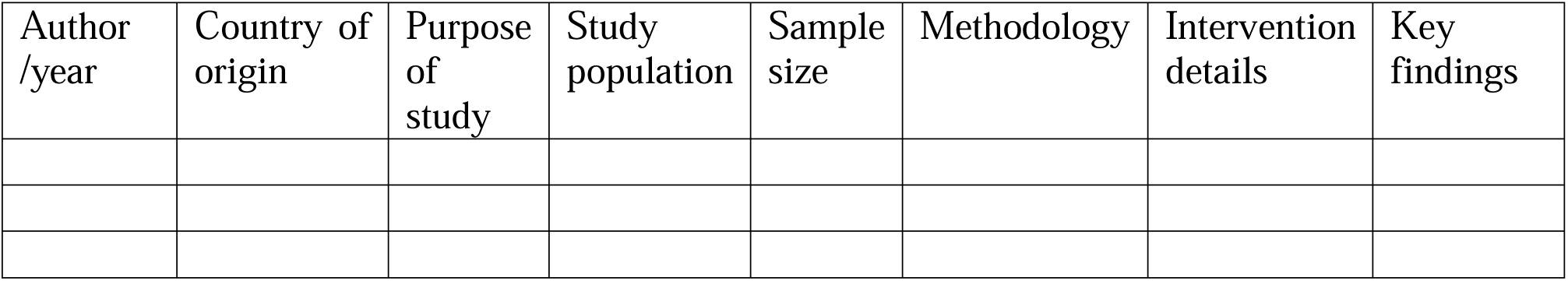
Sample data extraction sheet.

### Stage 5: Collecting, summarizing and reporting the results

The results will be summarized using multiple methods: numeric, narrative and thematic approaches. The studies will be summarized under the headings; distribution by geographical location, journals, year of publication, number of participants. Others will include; number of assessments, inclusion and exclusion criteria, interventions, quality assessment and analysis of themes.

## Quality assessment of data

Quality assessment of the data is not part of the processes involved in a scoping review as compared to systematic review ^35^, which is seen as a limitation according to Arksey and O’Malley^33^. The JBI emphasizes that the non-inclusion of quality assessment in a scoping review could limit the practical usefulness^34^. Although some authors have argued whether or not if a quality assessment is needed in this type of study, no further recommendations have been made till date. The JBI Critical Appraisal Checklists will be used for assessing methodological quality.

## Conclusion

The scoping review of interventions for caregivers of children with DD will provide a detailed summary of the available evidence for the effectiveness of interventions in improving the well-being of caregivers.

## Data Availability

All data produced in the present work are contained in the manuscript

## Patient and public involvement

This scoping review will neither include the patient nor the public.

## Ethics and dissemination

As a scoping review, the aim is to synthesize the current breadth of knowledge on the interventions rolled out for caregivers of children with developmental disabilities, no ethics approval is required. After completing this scoping review, the results will be disseminated through publication in a peer-reviewed journal.

## Footnotes

### Contributors

DAA and NN initiated and designed the study. DAA and JNS will do the screening of abstract and full text with disagreement to be resolved by NN. All authors read and approved the final manuscript.

### Patient consent for publication

Not required.

## Acknowledgement

The University of Witwatersrand, Faculty of Health Sciences Library provided technical support.

